# Designing and evaluating a health system resilient to extreme weather events in rural Madagascar

**DOI:** 10.1101/2025.01.06.24319322

**Authors:** Michelle V. Evans, Elinambinina Rajaonarifara, Andres Garchitorena, Fianamirindra A. Ralaivavikoa, Paulea Eugenie Rahajatiana, Karen E. Finnegan, Laura Cordier, Luc Rakotonirina, Bénédicte Razafinjato, Tokiniaina M. Randrianjatovo, Christophe Révillion, Malazafeno Jocelyn Mbimbisoa, Matthew H. Bonds

## Abstract

Adapting health systems for climate change can lessen the negative impact of climate change on human health. Even when not targeting climate-health links explicitly, broad health system strengthening interventions (HSSi’s) can ensure that the health workforce, infrastructure, and networks are robust enough to respond to and recover from climate-driven shocks. We explored the ability of an HSSi in a rural health district of southeastern Madagascar to serve as a climate change adaptation in response to cyclone Batsirai in 2022. The HSSi provides support for programs and health system infrastructure while introducing enhanced protocols and prioritizing rapid, local learning and adaptation. We conducted interrupted time series analyses of eight indicators of infectious disease and health system performance to assess the impact of Batsirai on two zones of the HSSi. We then examined how traditional domains of HSS, such as physical and human resources, combined with less formal domains, such as collective values, influenced health system resilience during this time. We found that the majority of indicators were resilient to cyclone Batsirai, with only vaccination rates affected in the two months following the cyclone, particularly in the zone where the HSSi had only begun 8 months prior. Changes in long-term trends were rare, and, when observed, revealed a slight slowing of progress, but not a regression to historical levels. After re-establishing the road network and providing additional supplies through an emergency response, the health system was able to resume routine service delivery without further external input and health system indicators continued to improve. The agility and responsiveness of the health workforce was enabled by formalized protocols, a culture of flexibility, open communication and data-informed action. HSSi’s that are designed to encourage local adaptation may increase health systems’ resilience to extreme weather events, resulting in health systems better adapted to climate change overall.

**TEASER KEY MESSAGE:** By improving overall service availability and readiness and establishing collective values that prioritize local and rapid adaptation, broad health system strengthening initiatives can create climate-resilient health systems.

**KEY MESSAGES:**

- Following cyclone Batsirai, a strengthened district health system in Madagascar mitigated the impact of this climate-driven shock on health system functioning. This was due to collective values of patient care, innovation, and data-informed action, in addition to supporting health resources and infrastructure.
- Rapid restoration of the transportation network via food-for-work programs and other external sources of support helped primary care facilities reopen quickly and resume admitting patients. Such coordinated approaches across sectors are needed to support health system responses to climate-driven natural disasters, and restore routine functioning.

## BACKGROUND

Cyclone Batsirai made landfall on the east coast of Madagascar on February 5, 2022.(Fig. 1). The combination of flooding from heavy rains and high wind speeds of up to 230 km/h resulted in disaster: over 7,400 homes were destroyed, displacing 61,489 people and impacting 112,115 across four regions^1^. In addition, 1,203 public schools were destroyed and 53 healthcare facilities were damaged, with six completely destroyed ^1^. Some regions reported over 90% of fields being destroyed^1^. The annual cyclone season often involves multiple cyclones across the island, many with devastating effects. Climate change is predicted to increase the intensity of cyclones in Madagascar in the coming decades^2,3^, increasing the climate vulnerability of a country which already experiences higher rates of poverty, food insecurity and mortality than neighboring nations^4,5^.

**Figure 1.**
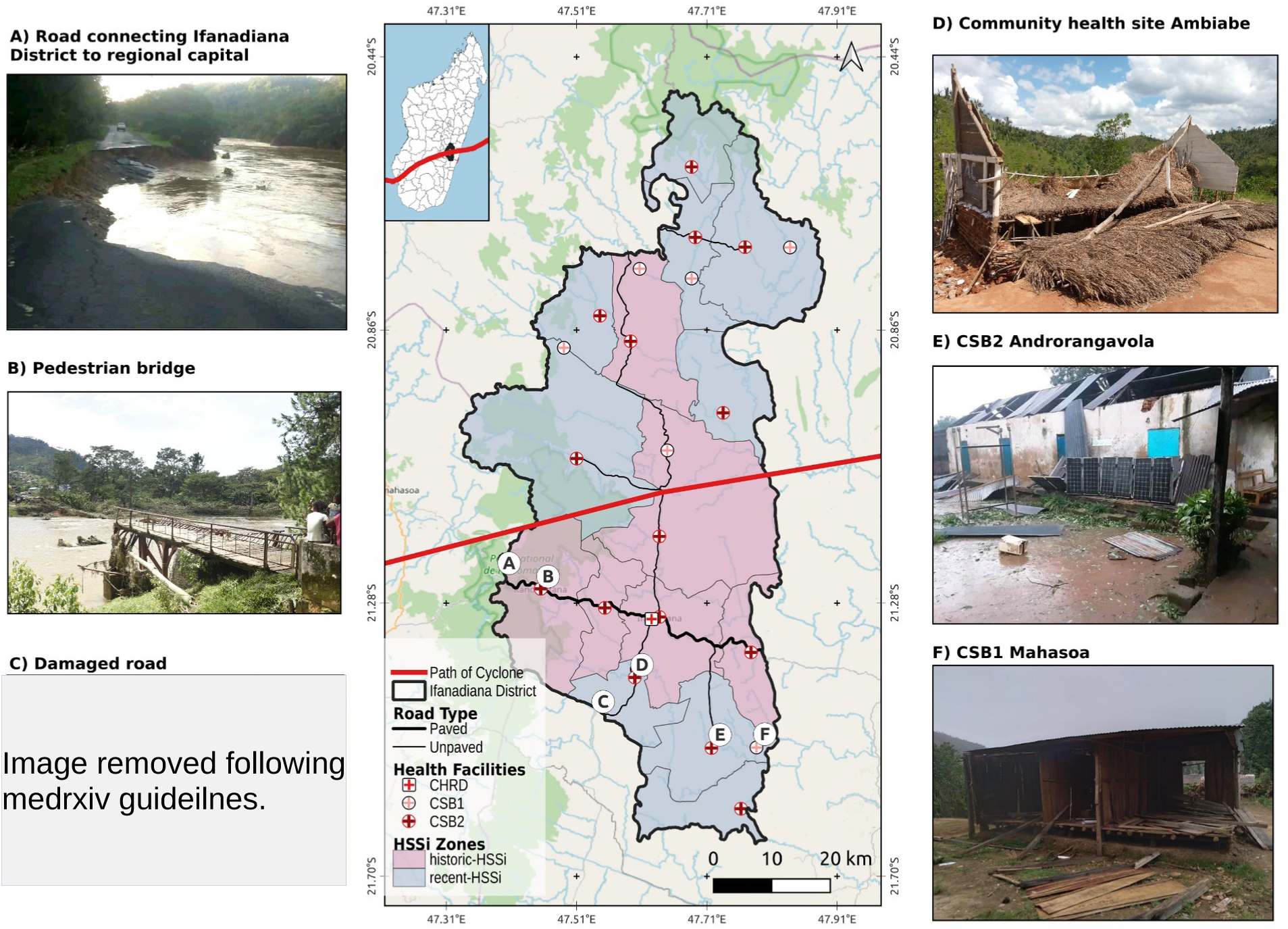
Cyclone Batsirai passed directly through Ifanadiana District (Vatovavy Region), Madagascar on February 5 2022, causing destruction of vital health and transportation infrastructure. The map illustrates the path of the center of Cyclone Batsirai through Ifanadiana District (inset shows the path in relation to Madagascar). The location of sites in the photos are identified via letters on the map, with a slight offset to reduce overlap. Shading represents historic-HSSi (beginning pre July 2021) and recent-HSSi zones (beginning post July 2021) in the district of Ifanadiana.

Madagascar is ranked 165 out of 185 on the Notre Dame Global Adaptation Initiative climate vulnerability index, given its high exposure as an island nation and the lack of adaptive infrastructure and governance, particularly the lack of regulation of government corruption and low agricultural capacity and innovation^6^. High climate vulnerability is associated with increased exposure to infectious disease, a limited ability to plan for and adapt to future scenarios, and more severe health consequences to extreme events^7^. Climate adaptations, such as resilient infrastructure or novel technologies and practices, can help mediate the impacts on population health. In addition, adapting health systems themselves to plan for and cope with the consequences of climate change can greatly mitigate its negative effects ^8,9^. Health system resiliency is one of the strategic priorities defined in Madagascar’s National Adaptation Plan, with a particular focus on early warning systems and adequate preparedness for emergencies^10^. In addition to climate-specific adaptations, strengthening the routine functioning of a health system can ensure that services are continuously available and have the flexibility to adapt to shifting population health needs as climate exposures change^11^.

Here, we explore the role of a health system strengthening intervention (HSSi) as a climate adaptation strategy via a case study of cyclone Batsirai in 2022 in southeastern Madagascar. We detail the health system programs related to cyclone preparation and response, and how the HSSi mediated the impact of the event. We then apply interrupted time series analyses to eight indicators of population health and health system functioning to compare zones of the district benefiting from a longer versus more recent support from the HSSi to evaluate how the cyclone impacted these indicators, and whether the effect of the HSSi differed across these zones.

### Adaptation Strategy

#### Study Population

Ifanadiana District is located in southeastern Madagascar, in a region identified as vulnerable to climate change^12^. The district is divided into 15 communes and 195 fokontany (the smallest administrative unit), with public health services overseen by the Bureau de Santé du District (BSD). The population of approximately 200,000 people is distributed across rural villages, although population density is higher at the center of each commune and along the main paved road, which runs east to west across the district (Fig. 1). Each commune is served by at least one primary health center (Centre de Base Niveau 2, CSB2), and six of the larger communes contain a second primary health center that provides more limited services (Centre de Base Niveau 1, CSB1) (Fig. 1). Each fokontany contains a community health site, staffed by two community health workers, who provide integrated community case management for children under five years old and limited maternal and reproductive health services^13^. The district hospital provides secondary care and is located in Ifanadiana, the district capital, with tertiary care provided via referral to the regional hospital 85km away in the regional capital of Fianarantsoa.

The health conditions in the district are characterized by high prevalence of infectious diseases and relatively low access to care, although coverage rates have risen since 2014 following the start of the HSSi^14^. A biannual longitudinal cohort study has been conducted since 2014 as part of the HSSi, enabling close monitoring of population health^15^. District-wide prevalence of diarrheal disease in children under-5 ranged from 9 to 19% between 2014-2018, with certain fokontany experiencing prevalence rates as high as 70%^16^. Malaria is endemic in the district and the prevalence in children under 15 years ranged from 0 - 80% across fokontany in 2021^17,18^. The 2023 survey found that nearly 50% of the population who reported illness in the prior month sought care, up from 25% in 2016^14,19^. Financial and geographic barriers are the most reported reasons for not seeking care^19^. Given the lack of transport infrastructure and the poor quality of roads, more than three quarters of the population live further than a 1 hour walk from a CSB and only 13% of the population lives within two hours from the district hospital^20,21^.

#### Health System Strengthening Intervention (HSSi)

Pivot, a non-governmental organization (NGO), began a partnership with the Madagascar Ministry of Public Health (MoPH) to build a district-level model health system in Ifanadiana District (Vatovavy) in 2014 via a health system strengthening intervention (HSSi). The partnership focuses on service delivery and integrated science, tackling issues of readiness, quality of care and social protection across community, primary, and secondary care levels of the public health system^22^. The intervention aims to increase health system functioning and resilience to serve a population faced with high rates of poverty in the context of a changing climate ^23^. While the HSSi does not include programs specifically targeting climate change, mitigating climate impacts on health is one of its goals, given the population’s vulnerability to extreme weather events.

The HSSi was based on similar initiatives undertaken as part of the multi-country *Population Health and Implementation Training* partnerships program^24^, particularly the partnership in Rwanda ^25^. Broadly, the intervention includes the six components of health system strengthening established by the World Health Organization^26^:service delivery; health workforce; information systems; medicines and supplies; financing; and governance. It focuses on the health district as the unit of study and employs multiple, continuous data streams to monitor and evaluate the intervention, drawing on an implementation science approach ^27^. The HSSi differs from similar interventions particularly in its approach to data and evaluation. In addition to operational data, a longitudinal cohort allows the collection and evaluation of population-level metrics and a multidisciplinary research team is embedded within the organization to produce high-quality science to both inform operations and contribute to broader scientific advancement. The HSSi began in 2014 in four communes of Ifanadiana, and expanded to two more communes before 2017, encompassing one third of the district’s population, referred to as the historic-HSSi zone in this study (Fig. 1). It was gradually expanded across the rest of the district by 2021, corresponding to the recent-HSSi zone (Fig. 1). The whole district was covered by the HSSi beginning in July 2021, and began expanding into two new districts in 2024. Details on the exact programs of the HSSi are provided in Table 1.

**Table 1.** Summary of HSSi implemented in Ifanadiana district beginning pre-July 2021 (historic-HSSi) and post-July 2021 (recent-HSSi) by WHO HSS component. Reproduced from Garchitorena et al.^14^

|  | **District Hospital (CHRD)** | **Primary care facility (CSB2)** | **Community health<sup>1</sup>** |
| --- | --- | --- | --- |
| **Service Delivery** | Network of 3 ambulances for referrals and emergency care; infrastructure renovations, provision of medical and non-medical equipment, including full laboratory capacity; social support for vulnerable patients | Infrastructure renovations, provision of medical and non-medical equipment; implementation of IMCI & malnutrition protocols for every child under 5, support for maternal health services | Construction of community health posts by community, with Pivot support; implementation of IMCI & malnutrition protocols for every child under |
| **Health workforce** | Staffing of health workers and non-clinical staff above MoPH norms; trainings for medical staff | Staffing of CSBs above MoPH norms; frequent trainings for medical staff | Training, coaching and monthly supervision of community health workers by mobile teams of trained nurses and midwives |
| **Health information systems** | Creation of a hospital-based M&E team to follow-up progress of activities; frequent facility readiness surveys | Joint MoPH-Pivot training and supervision to improve HMIS data quality | Joint MoPH-Pivot training to improve HMIS data quality ; support for use of electronic tools for data collection and decision making |
| **Medicines and supplies** | Supply chain management to reduce stock-outs, management of hospital pharmacy | Supply chain management, training, and reduction of stock-outs | Monthly provision of MNCH medicine stocks to CHWs and follow-up of medicine stock use |
| **Financing** | Cost of outpatient and inpatient care fully covered for referred patients (district hospital and tertiary care outside Ifanadiana) | Essential medicines and consumables provided free of charge to all patients | Cost of MNCH medicine stocks fully covered by Pivot; financial incentives to CHWs for stock management and attendance to supervisions |
| **Leadership and Governance** | Creation of a joint MoPH-Pivot executive committee for hospital management and transparency; subcommittees for specific projects | Close collaboration with district health managers for the planning and implementation of activities | Community engagement and participation |

<sup>1</sup>historic-HSSi only MNCH: maternal, newborn, and child health CHW: community health worker

Rapid, local adaptation is key to the HSSi. Service availability and readiness is addressed through context-specific infrastructure, such as signal-boosting telecommunications antennas in remote villages, and customized protocols to meet patient needs, such as the pilot of a proactive community care program^13^. In addition to the formal program design, local adaptation is also encouraged on an everyday basis through collective values that are established within the partnership, including ‘care for the patient’, ‘solidarity’, ‘curiosity’, ‘bias towards action’ and ‘embracing complexity’^28^. The collective values of the HSSi partnership are similar to the idea of the *intangible software* of a health system in complex adaptive systems approaches in that it characterizes the relationships, communication practices, and norms ^29–31^. To better respond to patient needs, the HSSi encourages the health district to be dynamic, enabled by a rigorous system of program monitoring and evaluation that tracks several hundred indicators of population health and health system functioning, as well as close partnerships with patients, care providers, and community leaders. For example, when data revealed significantly lower rates of maternal health service coverage in more geographically isolated communities, new programs were created to better engage community health workers (CHWs) and traditional birth attendants in maternal health^22^. In addition, maternal waiting homes were constructed to provide lodging for expectant mothers who live further from the CSB. The HSSi is therefore the combination of the resource and programmatic support relevant to the WHO HSS components and the more abstract set of collective values that enable rapid, local adaptation.

## APPROACH

We used mixed-methods to document if and how the HSSi served as a form of climate-change adaptation in the context of cyclone Batsirai. We analyzed the difference in multiple health system indicators via time series analyses, used satellite imagery to assess the impact of Batsirai on transport infrastructure, and relied on archival records and targeted discussions to provide context and essential details about how the health system performed during that period. Archival documents from the time before and after landfall by Batsirai included meeting minutes from internal daily emergency meetings, daily and weekly reports on health system infrastructure collected by the MoPH and Pivot, daily reports of the needs of displaced persons at all temporary shelters across the District, and internal and external evaluation reports of the response. Co-authors reviewed documents and jointly established a consensus of HSSi activities at that time.

We conducted statistical analyses of the monthly time series of select indicators from January 2021 to December 2023 from the district health information system relevant to infectious disease and health system performance to compare the historic- and recent-HSSi zones. These zones were determined by how long they had benefited from the HSSi, which was implemented gradually across the district. The historic-HSSi zone corresponds to the six communes benefiting from the HSSi before July 2021, and the recent-HSSi zone corresponds to the nine communes where the HSSi began in July 2021 (Fig. 1). We therefore considered two separate geographic zones of the district in the analysis to better compare the impact of the historic-HSSi and the recent-HSSi on the health system’s ability to adapt to and recover from the cyclone (Table 1, Figure 1).

We identified eight indicators of infectious disease and health system performance: reported malaria case rates; malaria rapid detection test (RDT) positivity rates; reported diarrheal disease case rates; the proportion of consultations diagnosed with diarrhea; outpatient consultation rates; referral rates; annual childhood routine vaccination coverage (first dose of diphtheria tetanus toxoid and pertussis (DTP3)); and the availability of UHC-tracer medicines. Monthly data on each indicator were collected by Pivot’s Monitoring and Evaluation team at the CSB level from January 2021 - December 2023. We applied a multi-group interrupted time series analysis to each indicator via a generalized linear mixed-model that controlled for long-term and seasonal trends in the data. The HSSi zone, the cyclone period (defined as February and March 2022), and the interaction between the two were included as fixed effects while the year, commune, and month of year were included as random effects. RDT positivity rates and the proportion of consultations diagnosed with diarrhea were modeled via Beta distributions with a log-link. The availability of UHC-tracer medicines was modeled via a Gaussian distribution and a log-link. Childhood vaccination coverage was estimated as an annualized percentage, rounded to the nearest percent, and modeled via a negative binomial distribution and log-link. All other indicators were transformed to a rate per 10,000 individuals and modeled via a negative binomial distribution and a log-link. All models were fit via the glmmTMB package^32^ and model residuals were tested for uniformity and over-dispersion via the DHARMa package^33^ in R v. 4.4.0^34^. Rate ratios (e.g. the difference in an indicator between two groups) were calculated for each indicator to facilitate comparison across indicators.

Because cyclone Batsirai arrived during an on-going HSSi, we undertook a supplementary analysis using longer-term data to explore whether cyclone Batsirai disrupted multi-year trends and negatively impacted the ability of the HSSi to improve the health system. This included the indicators mentioned above, with the addition of consultation rates of CHWs and severe malnutrition case intake rates at CSB2s, collected monthly from January 2017 - December 2023. The interrupted time series analysis included time in months since January 2017, the HSSi zone, a binary variable representing the post-cyclone period, months since the arrival of Batsirai (February 2022), the time since the intervention in the recent-HSSi zone, the interaction between time and the HSSi zone, and the interaction between the post-cyclone period and the HSSI zone as fixed effects. We included commune and month of the year as random effects and an AR(1) autoregressive temporal model for each commune to control for temporal autocorrelation in the data. Because the recent intervention began only 8 months before the arrival of Batsirai, we were unable to include an interaction between the HSSi zone and the months since Batsirai without introducing singularity into the model. The results of these models are therefore primarily to explore how Batsirai disrupted long-term trends in each HSSi zone, not to statistically test the difference in this disruption between the two zones. Indicators were transformed and fit as described above for the short-term analysis.

We used satellite imagery to identify areas impacted by cyclone-related flooding, particularly surrounding road networks between CSBs and the district capital, where the District health office is based. We collected Sentinel-1 radar imagery at the Analysis Ready Data level from the nearest satellite passage dates before (January 27 2022) and after (February 8 2022) cyclone Batsirai. Analysis Ready imagery was accessed at Digital Earth Africa (https://www.digitalearthafrica.org/), and included radiometric terrain correction and normalization of backscatter values. We calculated the normalized difference ratio between the two radar images on the VV and VH polarization independently and applied a threshold of −0.75 to identify pixels that were flooded in the post-cyclone image, following Johary et al.^35^. We combined this information with a road network of Ifanadiana collected as part of a community cartography effort that contains more than 20,000 km of roads and footpaths in the district^20^. We identified the optimal route between each CSB and the district capital along roads that are passable by motorcycle or car, which represent the route used to distribute medicines and supplies and refer patients to the District hospital. We then noted sections of each route that were within 50m or 100m of flooded areas to identify disruptions to the transportation network post-cyclone.

Use of aggregate monthly consultation counts for this study was authorized by the Madagascar Ministry of Health and the Regional Health Direction of Vatovavy Region. It was deemed non-human subjects research by Harvard University’s Institutional Review Board.

## FINDINGS

### Health system preparations

Preparation for the arrival of Batsirai began a week before its predicted arrival, and included all levels of the health system. A multi-sectoral meeting was held in Antananarivo, the national capital, between partners at the MoPH, the National Office for Risk and Disaster Management (*Bureau Nationale de Gestion des Risques et des Catastrophes*), internal technical and financial partners (e.g. UNICEF, Médecins sans Frontières), and local and regional NGOs, including Pivot. The goal of this meeting was to plan for the response and prepare financial and physical resources that might be required. A preexisting Memorandum of Understanding (which details the roles and responsibilities of the MoPH/Pivot collaboration), and relationships that developed from a history of regularly-held planning meetings as part of the HSSi, ensured district representatives were active collaborators at the central and local levels. As part of this effort, an emergency stock of medicine established as part of the HSSi was drawn on to supply the mobile health clinics that would serve displaced residents of Ifanadiana. In addition, the alternative supply of medicine was available to limit stock disruptions at CSBs. In the period following the cyclone, CSBs witnessed an increase in the availability of UHC-tracer medicines compared to the three-year average (Fig. 2, Table 2), suggesting this was a success.

**Figure 2.**
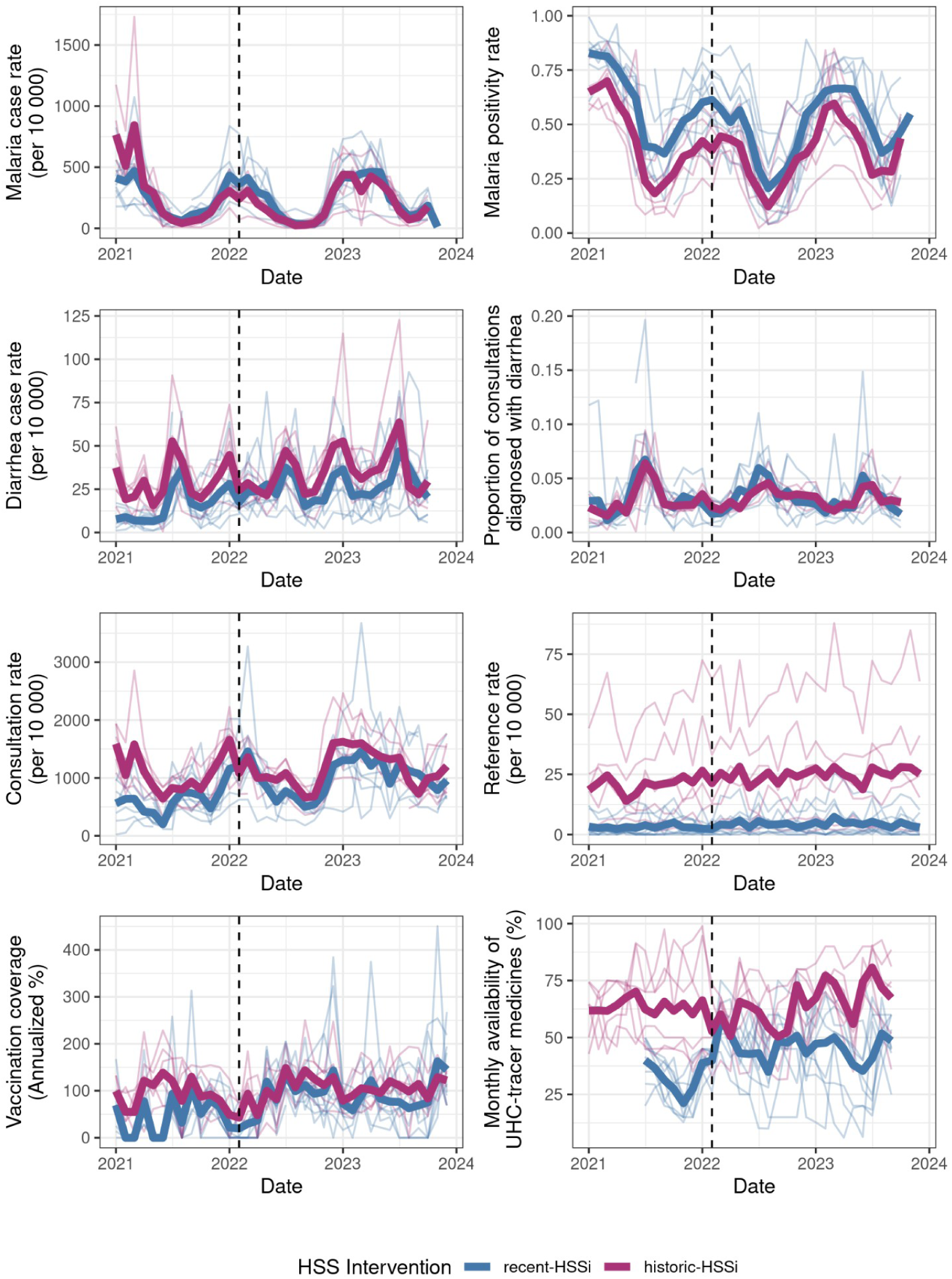
Monthly time series of eight indicators of population health and health system functioning from January 2021 -December 2023. The dashed line represents February 2022, the month when cyclone Batsirai arrived. Fine lines represent commune-level means while the bolder lines represent the mean by HSSi-zone (historic vs. recent). One outlier value has been removed from the graph of vaccination coverage to aid with visualization. Historic-HSSi refers to the zone benefitting from the intervention prior to July 2021 and recent-HSSi represents the zone where the intervention began in July 2021.

**Table 2.** Results of GLMMs estimating the association between the HSSi-zone, the post-cyclone period, and the interaction between the two on eight indicators. Rate ratios represent the ratio between the two groups (i.e. historic vs. recent HSSi, post-cyclone period vs. long term average, and impact of cyclone on historic vs. recent HSSi) for each indicator. 95% CI are shown in parentheses.

| Indicator | Historic-HSSi |  | Post-Cyclone Period |  | Historic-HSSi x Post-cyclone |  |
| --- | --- | --- | --- | --- | --- | --- |
|  | Rate Ratio | P-value | Rate Ratio | P-value | Rate Ratio | P-value |
| **Malaria** |  |  |  |  |  |  |
| Reported case rate | 0.86 (0.6 - 1.25) | 0.434 | 1.32 (0.97 - 1.8) | 0.077 | 0.76 (0.51 - 1.15) | 0.195 |
| Malaria RDT positivity rate | 0.53 (0.33 - 0.83) | 0.005 | 0.8 (0.6 - 1.07) | 0.138 | 0.92 (0.63 - 1.34) | 0.648 |
| **Diarrhea** |  |  |  |  |  |  |
| Reported case rate | 1.55 (1.16 - 2.05) | 0.003 | 1.15 (0.86 - 1.53) | 0.344 | 0.9 (0.63 - 1.28) | 0.554 |
| Proportion of positive consultations | 1.07 (0.83 - 1.38) | 0.589 | 0.79 (0.56 - 1.13) | 0.196 | 1.29 (0.83 - 2) | 0.259 |
| **Consultation rate** | 1.39 (1.08 - 1.78) | 0.01 | 1.48 (1.14 - 1.91) | 0.003 | 0.65 (0.46 - 0.91) | 0.012 |
| **Referral rate** | 6.72 (2.03 - 22.27) | 0.002 | 0.76 (0.57 - 1.03) | 0.079 | 1.18 (0.85 - 1.65) | 0.316 |
| **DTP3<sup>1</sup> vaccination coverage** | 1.51 (1.15 - 1.99) | 0.003 | 0.37 (0.18 - 0.77) | 0.008 | 1.83 (0.72 - 4.65) | 0.207 |
| **UHC-tracer availability** | 1.59 (1.37-1.83) | 0.000 | 1.19 (1.02-1.38) | 0.022 | 0.74 (0.6-0.91) | 0.004 |

<sup>1</sup> Diphtheria tetanus toxoid and pertussis, indicator representing routine childhood immunization coverage

Within the district, staff were mobilized at both the CSBs and MoPH and Pivot central offices to prepare for the response. Teams were formed following existing operational roles to leverage existing expertise and relationships and ensure a dynamic response. For example, to enable the distribution of over 1000 food and hygiene kits before February 5, the supply team organized the purchase and creation of the kits, the transportation team organized vehicles for their distribution, and the M&E team closely tracked the arrival of displaced persons and kit distributions to plan for future deliveries. These teams were a part of Pivot’s operational workforce, in addition to MoPH staff, and had experience in organizing and improving supply chain management in partnership with MoPH staff. With a well-trained workforce and a history of collaboration, the operational teams were able to rapidly adjust to this new responsibility. The distribution of emergency kits continued throughout the response, ultimately distributing 2,361 kits across the district.

Medical and operational staff were deployed to more isolated regions of the district prior to February 5, and remained there during the response to support community-based responses. Not only did this ensure close monitoring of the populations’ needs and health, but the physical placement of personnel at each site enabled an approach that centered on, and was tailored to, the needs of the community and patients. Health staff accompanied village leaders to conduct education campaigns about the coming cyclone, distribute food kits, and encourage those in vulnerable housing to relocate to temporary shelters. This collaboration was enabled by long-standing informal and formal relationships between local communities and health staff, encouraged by the HSSi’s collective values of ‘solidarity’ and ‘care for the patient’. These preparations, particularly the organization of temporary shelters, were a success. Because of outreach to identified at-risk households, families were supported to move to temporary lodging. As a result, despite homes being destroyed and landslides recorded, no deaths were directly attributed to cyclone Batsirai in Ifanadiana district.

### Immediate health system response

The immediate response focused on supporting those who were directly impacted by the cyclone and ensuring that routine care was able to continue, particularly for climate-sensitive infectious diseases, such as malaria and diarrheal disease^36^. This was done via the deployment of mobile clinics which provided daily, on-site health and social consultations at temporary shelters and the quick re-opening of CSBs. Mobile clinics were staffed by MoPH clinicians, and medical supplies were drawn from standard MoPH stock and the HSSi emergency fund. Cases were reported to the District office daily via text message, and eventually integrated into the standard monthly reports of each CSB. This was facilitated by the HSSi’s focus on communication, which included the establishment of alternative channels of communication via satellite phones and creation of *ad hoc* committees and response teams. There were outbreaks of malaria in two communes, but these were quickly reported to the District office during the daily report, and emergency teams were deployed to respond. Neither malaria nor diarrheal disease saw an increase in reported case rates or positivity rates following the cyclone (Table 2, Fig. 2).

Heavy rains and winds significantly disrupted road accessibility across the district, which hampered the distribution of supplies and the movement of patients. Radar imagery revealed that the primary transportation routes linking CSBs to the District hospital remained impacted by cyclone-related flooding on February 8, 3 days post-Batsirai, and internal reports cite that only 5 of 15 communes were accessible on this date (Fig. 3). The primary paved road in the district was severed on the route towards the regional capital of Fianarantsoa in Vohiparara, cutting off the region from road transport with the rest of the country for several days (Fig. 1A, Fig. 3). A relay ambulance system, similar to that already in use in the HSSi program, was established to connect ambulance routes on either side of this road block. This system served to deliver supplies and medical workers to the district and to deliver patients in need of tertiary care to the hospital. A “work-for-food” program was put in place to re-establish key transport routes that had been destroyed, including removing trees from roads and rebuilding a bridge (Fig. 1B). This program was unique in that it brought together partners who traditionally did not collaborate, including the military police, the local government, and local actors working on a range of issues from biodiversity protection, education and health. This collaboration was emblematic of the sense of solidarity felt among partners at this time of crisis. Although technically outside the scope of health service delivery, repairing the road network immediately removed a significant barrier to accessing health facilities, with positive effects on consultation rates, and contributed to food security during this time. Both zones witnessed an increase in consultation rates following Batsirai, relative to long-term trends (Table 2, Fig. 2).

**Figure 3.**
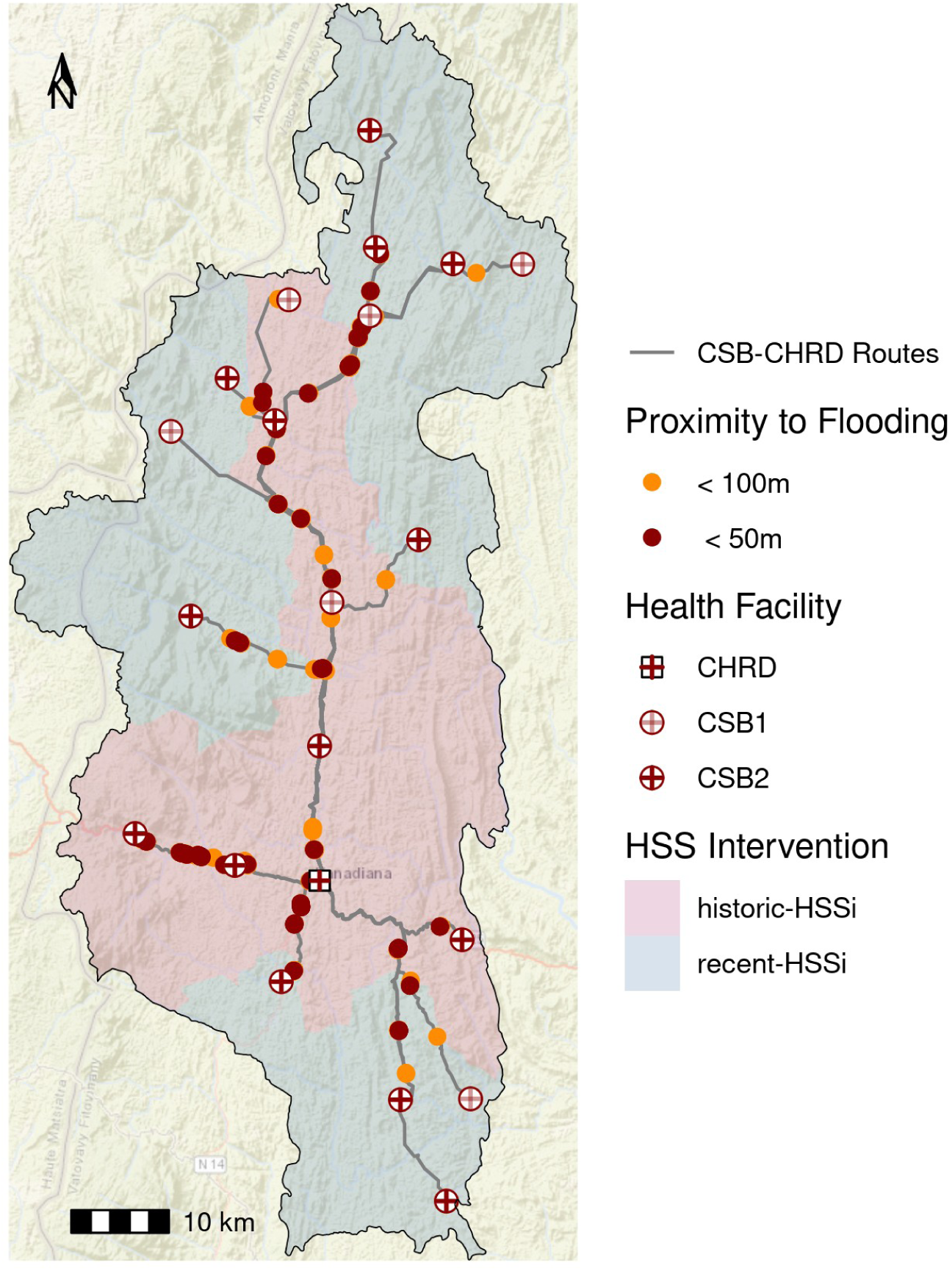
The motorized vehicle transportation network remained significantly impacted by flooding 3 days post-cyclone. The motorized vehicle network connecting CSBs to the District hospital (CHRD) on February 8 2022. Portions of the road network in close proximity to flooding are highlighted. Base Map: ESRI World Street Map.

Health facility infrastructure was similarly impacted. At the District hospital, the temporary ward that was constructed for COVID-19 patients was blown away. Three months after the outbreak of the Omicron variant in nearby South Africa, this represented a dangerous confluence of crises. The loss of roofs and solar panels led to loss of electricity and consequent loss of vaccination cold chains at three CSBs. Although clinic heads worked with communities to move refrigerators to more secure locations in private houses, vaccination coverage at CSBs decreased in the period following the cyclone (Table 2, Fig. 2). This decrease may have been due to the destruction of stock following the cyclone or the lack of reinforcement of vaccination programs at that time. Vaccination data is also of particularly low quality during this time period, with 20 of 21 CSBs in the district missing data on polio vaccination coverage in February 2022, suggesting preventative care programs, such as vaccination, were particularly impacted by cyclone Batsirai.

### Long-term impact of cyclone Batsirai

Cyclone Batsirai arrived in the midst of an on-going HSSi, and there was concern among partners that it would disrupt the intervention itself, rolling back advances that had been made since 2014^14^. In fact, a sense of ownership of the success of the HSSi, and an associated responsibility for it, was one of the motivations cited for such a strong disaster response by personnel. Our supplementary analysis found little evidence for a significant impact of Batsirai on long-term trends in indicators (Fig. 4, Table S1). Of the indicators with the most positive trends pre-Batsirai - notably consultation rates, malaria case rates, and referral rates - only the increase in consultation rates slowed post-February 2022. However, it is difficult to statistically disentangle this from the expected plateauing of consultation rates over time (Fig. 4). Those indicators which were most impacted in the short-term by Batsirai, particularly vaccination coverage and the availability of UHC-tracer medications, have slightly increased in the years since Batsirai, suggesting they are not just recovering from the disruption, but continuing to improve (Table S1, Fig. 4).

**Figure 4.**
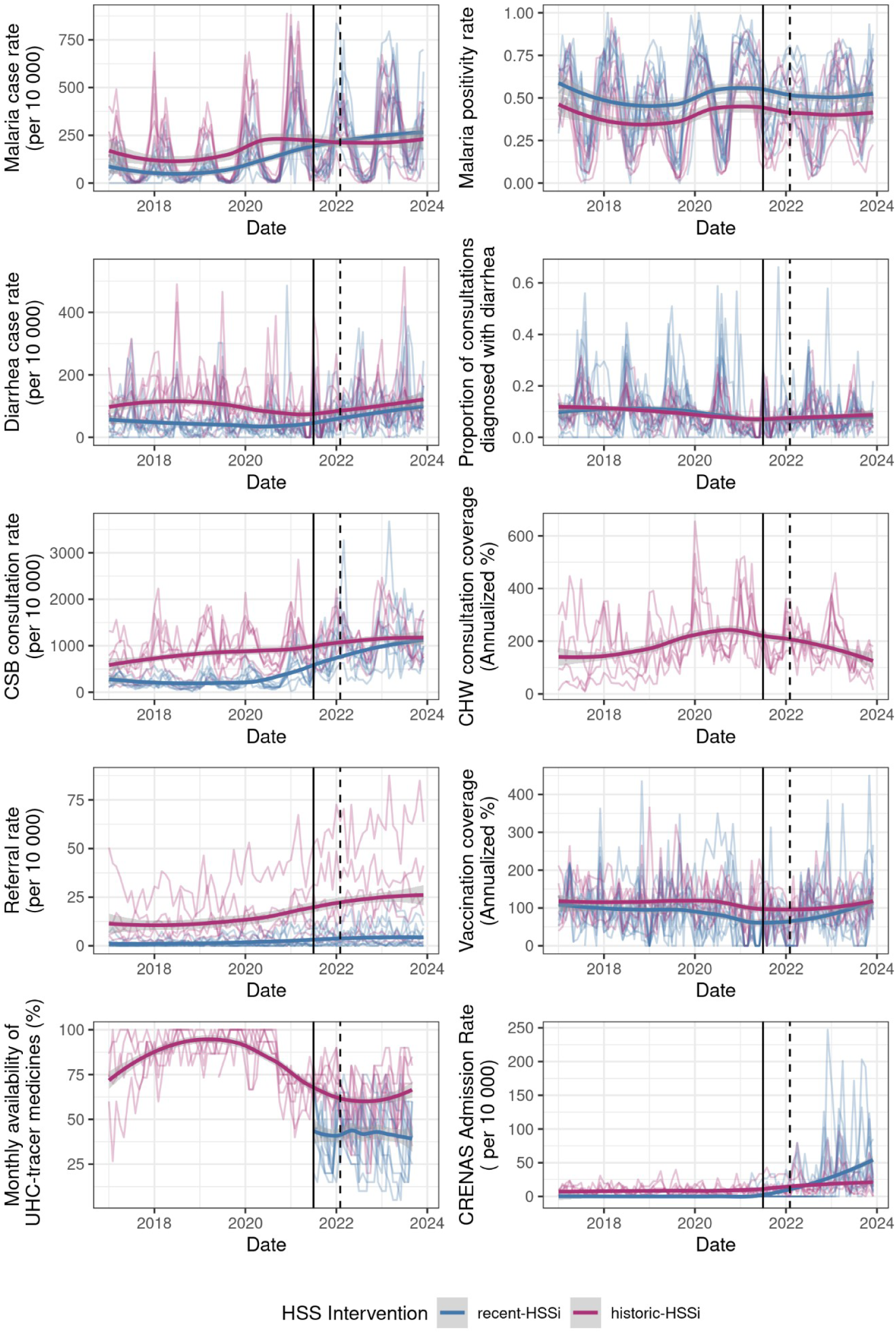
Long-term trends in indicators from January 2017 - December 2023. Indicators are plotted at the commune level (in lighter shaded lines), with loess curves added for each intervention zone to aid with visualization of long-term trends. One upper outlier each of malaria case rates, vaccination coverage, and malnutrition intake rates have been removed to aid with visualization.

Cyclones can impact food security in both the short- and long-term by destroying crops, disrupting supply chains, and increasing prices^37^. To manage short-term food insecurity, food kits were distributed at temporary sites and within communities before and immediately following the cyclone. Very few households in Ifanadiana have refrigerators or the capacity to store food, making them extremely vulnerable to disruptions to food supplies. While the distribution of food kits may fall outside of a conventional health sector activity, it was recognized as an immediate need of the population to prevent a larger health crisis, and HSSi programs were able to adapt to serve this need. The distribution of food kits in the weeks following Batsirai helped reduce the risk of immediate starvation after the cyclone. However, in some communes, nearly 90% of rice and manioc fields, the population’s primary food sources, were destroyed by Batsirai, increasing the risk of long-term food insecurity. Madagascar was already in the midst of a malnutrition crisis^38^, and Batsirai further aggravated this. Severe malnutrition case intake of children across the district increased from an average of 32 monthly admissions from January 2021 - January 2022 to an average of 92 from April 2022-December 2023 (Fig. 4, Table S1).

## DISCUSSION

Building resilience to climate-related shocks is an increasingly important role of health system strengthening programs. We used a mixed-methods approach to study the impact of cyclone Batsirai in 2022 on a rural health district in southeastern Madagascar and the HSSi’s ability to mitigate this impact. We examined the activities during the preparation for, immediate response to, and long-term follow-up of cyclone Batsirai. Routine care continued relatively undisrupted, facilitated by the deployment of mobile health clinics and the rapid re-establishment of transport networks, and infectious disease outbreaks were minimal. Even though the HSSi was not explicitly designed for climate-driven shocks, its combination of resource and programmatic support and the cultivation of an environment of rapid, local adaptation in the partnership contributed to the resilience of the health district at that time.

A population’s vulnerability to climate-driven health hazards is the result of a complex interaction of social, economic, institutional and environmental determinants. In Ifanadiana, differences in socio-economic status are some of the primary determinants of exposure to health hazards, access to care, and resulting health outcomes ^16,39,40^. The primary role of the health system is to provide service delivery to all patients. However, the HSSi has also targeted the historic inequality in healthcare access across socio-economic levels via fee reimbursement and social programs, resulting in a reduction in economic inequality in care-seeking and maternal health indicators since 2014^14^. Similarly, the patient-centered approach used during the cyclone response addressed patients’ needs across multiple dimensions of vulnerability, not just health. Poorer households are disproportionately impacted by natural disasters^41^, and the HSSi’s existing focus on health equity remained during the response period. The distribution of emergency food and hygiene kits and availability of social workers at temporary sites aimed to address patients’ most urgent needs outside of curative care. The deployment of mobile clinics to temporary shelters also ensured that those most vulnerable at the time would receive healthcare, and helped prevent a multiplicative effect of combined crises on patients’ health.

The overall post-cyclone response in the district was viewed as successful: routine care was provided continuously, the few disease outbreaks that occurred were contained, and no deaths were attributed to the cyclone. In addition, indicators of health system functioning continued to improve, evidence that the HSSi programs themselves were resilient to this disruption. Although somewhat hidden by the rapid rebound seen in indicators of health system functioning, the effects of Batsirai on the population’s standard of living and infrastructure were severe and long-lasting. As of September 2024, many government buildings still lack roofs or walls, some have been abandoned altogether, and several health facilities continue to work out of what was intended to be temporary tents. These infrastructural challenges have not prevented the provision of routine care, but they do highlight that recovery is on-going over two years later. This impact manifests most clearly in childhood malnutrition. In the months following the cyclone, malnutrition cases increased by over 300%. Data collection following the cyclone noted significant damage to crops, but the immediate response did not include a program to prepare for the indirect impact this could later have on food security. However, the HSSi’s approach of data-informed programs has since enabled the rapid creation of a new program in the district to pro-actively search children at risk of moderate malnutrition, in direct response to the observed increase in severe cases.

HSS is broadly viewed as the combination of increasing investment in resources, in both the short and long-term, and structural changes to policy and behavior, with a focus on impact at the system-level that cuts across the WHO HSS components^42,43^. In the case of natural disasters, both approaches are needed, as was seen during Batsirai. The HSSi supported the population immediately post-cyclone via its involvement in the donation of and distribution of food kits and food-for-work programs re-establishing road networks. Both of these involved the input of external resources via an unrestricted, emergency fund. Once these major obstacles were removed, the health system was able to quickly return to a normal routine and absorb the higher consultation rate seen post-cyclone, while preventing an increase in disease positivity rates. While there is a concern that HSS approaches that rely heavily on top-down, external assistance are less resilient and equitable than bottom-up, transformative approaches^44–46^, external inputs, whether from international donors or simply in addition to routine health system funding, may be necessary during times of natural disasters. Multi-sectoral approaches that include local disaster management and transportation offices, for example, are needed to support health systems in these extreme circumstances. Further, in regions where natural disasters are expected to become more routine, as in the case of Madagascar, disaster management should be considered in strengthening initiatives, and not regarded as rare events.

More than two years after cyclone Batsirai, the HSSi is expanding to cover the Vatovavy region, including three districts and a population of 1 million people. The new districts contain over 100 km of coastline and are particularly vulnerable to flooding given their low elevation. However, the majority of the HSSi has not substantially changed to adapt to this increase in climate vulnerability. Rather, the HSSi is being expanded following “adaptation with robustness”^47^, with a focus on fully staffing and training the health workforce while removing point-of-care user fees for pregnant women and children under-5. Indeed, the initial HSSi in Ifanadiana did not include climate- or cyclone-specific adaptations, but rather increased the system’s ability to adapt to and recover from shock by focusing on its “everyday resilience”^31,48^. Everyday resilience presupposes that care providers face frequent disturbances such as political instability, changing policies, and disruptions to medical supplies. By equipping care providers and systems to respond to these more common shocks, they are better able to respond to rarer, more extreme shocks. By applying a comprehensive suite of programs to the new districts, rather than focusing on their climate vulnerability specifically, and continuing to encourage a culture of local adaptation, the HSSi aims to increase climate resilience indirectly and not at the expense of patients’ overall health. The findings from this case study suggest that this approach is effective.

The HSSi described in this case study was not designed explicitly as a climate change adaptation. However, in its efforts to build everyday resilience through the support of health infrastructure and supplies and the cultivation of an environment that encourages local adaptation, the HSSi was surprisingly resilient to the arrival of cyclone Batsirai, one of the strongest tropical cyclones to reach the region in the past decade. This raises the question of how climate change should be considered in the design of HSSi’s and the larger roles of HSSi’s as climate change adaptations. In this case study, many of the characteristics of the HSSi that helped the system cope with common disruptions, particularly those not directly related to the WHO HSS programmatic components, were equally beneficial during an extreme weather event. If HSS is to serve a larger role in climate change adaptation, attention must be paid to these more emergent properties of health systems undergoing strengthening.

## Data Availability

All data produced in the present study are available upon reasonable request to the authors.

## AUTHOR CONTRIBUTIONS

Conceptualization - MVE, AG, FAR, KEF, LC, MHB. Methodology - MVE, ER, AG, FAR, TMR, CR, MHB. Software - MVE, ER, TMR, CR. Validation - all authors. Formal analysis -. MVE, ER, TMR. Investigation - All authors. Data curation - MVE, ER, FAR, PER, LC, LR, BR, TMR, CR, MJM. Initial Draft - MVE, KEF, MHB. Reviewing and Editing - All authors. Visualization - MVE, ER, TMR. Supervision - AG, KEF, LF, LR, BR, MHB. Project administration - MVE, KEF. Funding acquisition - MVE, FAR, KEF, LC, MHB.

## ACKNOWLEDGEMENTS

This manuscript is a part of a supplement titled, Lessons from the field: Case studies to advance research on climate adaptation strategies and their impact on public health. This writing project was supported by the National Institutes of Health (NIH) Climate Change and Health Initiative (https://climateandhealth.nih.gov) and coordinated by the Center for Global Health Studies at the Fogarty International Center of NIH. The activity was led by a steering committee of global experts on health and climate change. More information is available at https://go.nih.gov/ClimateAdaptationStudies. The authors would like to thank Ifanadiana District Ministry of Health staff for their response efforts following cyclone Batsirai and their continued work for the health of the people of Ifanadiana.

## FUNDING STATEMENT

This project was funded by a grant from the National Institute of Health Fogarty International Center.

## DISCLAIMER

NA

## COMPETING INTERESTS

Some authors are current or former employees of institutions discussed in this article, including the non-governmental organization PIVOT. These affiliations are explicitly listed in the article.

## Notes

### Funding Statement

This study was funded by the National Institutes of Health (NIH) Climate Change and Health Initiative and coordinated by the Center for Global Health Studies at the Fogarty International Center of NIH.

### Author Declarations

Use of aggregate monthly consultation counts for this study was authorized by the Madagascar Ministry of Health and the Regional Health Office of Vatovavy Region. It was deemed non-human subjects research by Harvard University's Institutional Review Board.

