## Supplementary material for "Designing and evaluating a health system resilient to extreme weather events in rural Madagascar": Table S1

**Table S1.** Results of GLMM models evaluating impact of Batsirai on long-term trends in indicators. UHC-tracer availability and consultation rates at CHWs were not complete enough in the recent-HSSi zone to compare between zones. Coefficients represent either rate ratios or exponentiated average monthly change (trends). 95% CI are shown in parentheses and those CI which do not overlap 1 are bolded.

| | Trend (slope over time) | historic-HSSi | Trend x historic-HSSi | Change in trend post-intervention (recent-HSSi only) | Change in intercept post-cyclone | Change in trend post-cyclone | Post-cyclone intercept x historic-HSSi |
|---|---|---|---|---|---|---|---|
| Malaria case rate | **1.04 (1.03 - 1.05)** | **3.33 (1.79 - 6.17)** | **0.98 (0.96 - 0.99)** | 0.94 (0.85 - 1.04) | 0.92 (0.6 - 1.39) | 1.07 (0.96 - 1.19) | 0.73 (0.4 - 1.33) |
| Malaria RDT positivity rate | 1.01 (1.01 - 1.02) | 0.64 (0.37 - 1.1) | 0.99 (0.98 - 1) | **0.86 (0.8 - 0.93)** | 0.89 (0.61 - 1.3) | 1.19 (1.1 - 1.29) | 0.68 (0.4 - 1.14) |
| Diarrhea case rate | 0.99 (0.99 - 1) | 1.83 (1.03 - 3.23) | 1 (0.99 - 1.02) | **1.13 (1.05 - 1.21)** | 1.01 (0.71 - 1.43) | **0.89 (0.83 - 0.95)** | 1.03 (0.65 - 1.66) |
| Proportion of consultations with diarrhea | **0.99 (0.98 - 0.99)** | 0.94 (0.67 - 1.33) | 1 (1 - 1.01) | 1.07 (0.99 - 1.15) | 1.16 (0.75 - 1.81) | 0.93 (0.86 - 1) | 1.43 (0.83 - 2.47) |
| Consultation rate at CSBs | **1.01 (1.01 - 1.02)** | **2.98 (2.01 - 4.41)** | 1 (0.99 - 1.01) | 1.17 (1.1 - 1.24) | 0.93 (0.7 - 1.24) | **0.86 (0.8 - 0.92)** | 0.77 (0.52 - 1.15) |
| Referral rate | **1.02 (1.01 - 1.04)** | **13.67 (3.72 - 50.32)** | 0.99 (0.98 - 1) | 1 (0.94 - 1.07) | 1.17 (0.84 - 1.62) | 0.97 (0.91 - 1.04) | 0.77 (0.53 - 1.11) |
| DTP3 vaccination coverage | 0.99 (0.99 - 1) | 1.06 (0.77 - 1.45) | 1 (1 - 1.01) | 0.98 (0.92 - 1.04) | 1.21 (0.87 - 1.7) | 1.04 (0.98 - 1.11) | 0.85 (0.55 - 1.3) |
| UHC-tracer availability | **0.98 (0.97 - 0.98)** | NA | NA | NA | **0.8 (0.66 - 0.98)** | **1.02 (1.01 - 1.04)** | NA |
| Consultation rate at CHWs | **1.02 (1.01 - 1.03)** | NA | NA | NA | 1.05 (0.82 - 1.36) | **0.96 (0.93 - 0.98)** | NA |
| Severe malnutrition intake rate | **1.07 (1.02 - 1.12)** | NA | NA | NA | 1.97 (1 - 3.87) | 1.06 (0.97 - 1.16) | NA |
